# Adverse Events and Baseline Psychological Distress Among Dementia Family Care Partners in the Tele-STELLA Study

**DOI:** 10.64898/2026.11.16.26368229

**Authors:** Christine McClure, Hailey Chatterton, Heather Franklin, Aimee Mooney, Allison Lindauer

## Abstract

**BACKGROUND:** Adverse event (AE) narratives may reveal physical and psychological challenges experienced by family care partners of people living with Alzheimer’s disease and related dementias.

**OBJECTIVE:** To characterize care partner AEs during the national Tele-STELLA study and explore differences in baseline psychological characteristics by AE category and subcategory.

**METHODS:** This descriptive, exploratory analysis included 43 AEs reported by 36 care partners. Narratives were classified as Physical, Psychological, or Random/Other. Baseline depressive symptoms, grief, and behavioral reactivity were compared between care partners with and without each AE type within this sample.

**RESULTS:** Physical events accounted for 48.8% of AEs, Psychological events for 39.5%, and Random/Other events for 11.6%. Infections predominated among Physical events; stress/distress accounted for half of the Psychological events and care-recipient aggression accounted for the other half of the Psychological events. Psychological AEs were associated with higher baseline grief and behavioral reactivity. Stress/distress-related AEs were associated with higher depressive symptoms and reactivity, whereas aggression-related AEs were associated with greater grief.

**CONCLUSIONS:** Care partner AE narratives captured heterogeneous physical and psychological experiences. Classification may complement conventional assessments of care partner health and distress. These exploratory findings do not establish causality or predictive validity and warrant prospective study.

**CLINICAL TRIAL REGISTRY NAME, URL, AND REGISTRATION NUMBER:** ClinicalTrials.gov, https://clinicaltrials.gov/study/NCT04627662, NCT04627662

## BACKGROUND & OBJECTIVES

Family care partners of people living with dementia have been described as invisible second patients,^1^ reflecting the substantial physical and psychological consequences that can accompany caring for a family member with Alzheimer’s disease and related dementias (ADRD). Physical consequences of caring for an individual with dementia can include increased morbidity, reduced engagement in health-promoting behaviors such as exercise and sleep, increased smoking and alcohol use, and greater use of prescription medications.^2–5^ Family care partners also report depression, anxiety, stress, guilt, social isolation, pre-death grief, and reduced self-efficacy.^1–3,6–10^ These challenges may be compounded by limited access to education and support for complex medical decision-making, management of behavioral symptoms, and other difficulties that may arise for ADRD care partners.^2,11,12^

In clinical research, adverse event (AE) monitoring is designed primarily to protect participant safety. The National Institute on Aging (NIA) defines an AE as “any untoward or unfavorable medical occurrence in a human study participant, including any abnormal sign (e.g., abnormal physical exam or laboratory finding), symptom, or disease, temporally associated with the participant’s involvement in the research, whether or not considered related to participation in the research.”^13^ Because an AE need not be caused by study participation, care partner AE narratives may capture conventional medical events as well as psychologically or relationally salient events occurring within the care experience. Care-recipient agitation and aggression are particularly relevant because they are among the most difficult and distressing dementia-related symptoms for care partners to manage.^14–16^ Family care partners may experience verbal, physical, or sexual aggression or other harmful behavior,^2,16,17^ often with limited confidence or training in how to prevent or manage these episodes.^2,11,15^ Such experiences can disrupt caregiving, increase care partner burden, and leave care partners feeling unsafe or violated.^16–18^

Despite extensive literature documenting care partner burden, psychological distress, and physical health consequences, care partner AEs themselves have received relatively little attention as an analytic outcome. AE data are generally collected for safety monitoring rather than examined as a potential source of information about care partner health, behavioral challenges, or periods of increased vulnerability. Free-text AE narratives collected during longitudinal care partner research may therefore provide a complementary perspective on the care partner experience that is not captured by conventional burden or symptom scales alone.

The primary objective of this descriptive and exploratory analysis was to characterize 43 care partner AEs reported during Tele-STELLA (Support via Technology: Living and Learning with Advancing Alzheimer’s Disease and Related Dementias), an NIA-funded telehealth intervention study for family care partners.^19^ Because AE reports were recorded as free-text narratives, we developed investigator-defined content categories to describe recurring clinical and care partner-related patterns. We also explored whether baseline care partner psychological characteristics differed according to subsequent AE category or subcategory.

## RESEARCH DESIGN & METHODS

### STUDY DESIGN & PARTICIPANTS

Data for this descriptive and exploratory analysis of care partner AEs were obtained from reports collected during Tele-STELLA (Support via Technology: Living and Learning with Advancing Alzheimer’s Disease and Related Dementias), an NIA-funded telehealth intervention designed to support family care partners of people living with ADRD (ClinicalTrials.gov NCT04627662).^19^ AE data were collected through weekly surveys in which care partners reported urgent care visits or hospitalizations involving themselves or the person with dementia, as well as through reports made during active Tele-STELLA sessions. After each AE, the Tele-STELLA coordinator contacted the care partner to collect required information and additional details about the event. The present analysis included care partners with one or more AEs reported during study participation. The Tele-STELLA study was approved by the Oregon Health & Science University Institutional Review Board (IRB #: 22288).

A detailed Data Safety Monitoring Plan (DSMP) was developed with the NIA Program Officer. This plan identified likely “expected” care partner events such as anxiety, back pain, and hypertension. A plan was outlined for a response from the Principal Investigator (PI) if the study team was concerned for the health or safety of care partners. All AEs were reported to the NIA in the quarterly reports. A Safety Officer oversaw all AEs, per the DSMP.^19,20^

### ADVERSE EVENT IDENTIFICATION AND CLASSIFICATION

Care partner AEs were extracted from the Tele-STELLA adverse event REDCap database and linked to the analytic dataset using participant study identification numbers. One investigator (CM) reviewed the narrative description of each AE and developed an initial classification framework through iterative review of the event summaries. Each AE was assigned to one primary category (Physical, Psychological, or Random/Other) and a clinically meaningful subcategory based on the content documented in the narrative.

Physical AEs comprised health or medical events such as infections, chronic disease exacerbations, acute or chronic pain, and falls; Psychological AEs included caregiving-related stress/distress, aggression from the person with dementia, and other psychological events; and Random/Other AEs reflected unrelated external events such as motor vehicle accidents, dog aggression, or being struck by a vehicle. Aggression-related events were classified according to the primary content documented in the AE narrative. Narratives about aggression varied in specificity, with some describing threatened physical harm and others documenting abuse without specifying its form. Because injury or medical consequences could not be inferred consistently, aggression-related events were retained in the Psychological category based on the event as documented rather than on potential injury risk. The framework was reviewed with the study PI (AL), and categories were refined through discussion until consensus was reached. Because individual care partners could experience more than one AE, a care partner could contribute events to more than one category or subcategory.

### MEASURES

Baseline demographic and care partner characteristics were summarized for care partners and care recipients. The 24-item Revised Memory and Behavior Problems Checklist (RMBPC) assesses standardized behavioral problems and care partner reactivity; total frequency and reactivity scores each range from 0 to 96.^16^ Baseline care partner psychological measures included RMBPC reactivity,^16^ the Marwit-Meuser Care Partner Grief Inventory (MM-CGI)^21^ to assess grief, and the Center for Epidemiologic Studies Depression Scale (CES-D)^22^ to assess depressive symptoms.

### STATISTICAL ANALYSES

Descriptive statistics were used to summarize care partner and care-recipient characteristics and the frequencies of care partner AE categories and subcategories. Continuous variables were summarized using means and standard deviations, and categorical variables using frequencies and percentages.

Exploratory analyses examined whether baseline care partner psychological characteristics differed according to AE type. Within the group of care partners who experienced at least one AE, independent-samples t-tests compared baseline CES-D, MM-CGI, and RMBPC reactivity scores between care partners who did and did not experience each specific AE category. Thus, the comparison group for a given category consisted of care partners who experienced at least one AE but did not experience an AE in that category. Mean differences, 95% confidence intervals (CIs), Hedges’ g, and p-values were calculated. Similar analyses were conducted for AE subcategories; subcategories experienced by fewer than three care partners were excluded because of unstable estimates.

Analyses were conducted using R version 4.1.2.^23^ Given the exploratory design, small event counts, and number of comparisons, findings were interpreted as hypothesis-generating, with emphasis on the direction and magnitude of observed differences rather than on isolated p-values.

## RESULTS

### CARE PARTNER CHARACTERISTICS

Baseline demographic and care partner characteristics are summarized in Table 1. Of the 188 enrolled care partners, 36 unique care partners (19%) experienced an AE. None of the baseline characteristics differed significantly between groups. The 43 care partner AEs included in this analysis were reported by these 36 unique care partners. Among the 36 care partners who experienced an AE, mean age was 68.7 years (SD = 9.9); most were female (72.2%), White (91.7%), non-Hispanic (97.2%), and living in urban areas (80.6%). Slightly more than half were spouses or partners of the person living with dementia (55.6%), and care partners had provided care for a mean of 3.9 years (SD = 3.9). Most reported sufficient financial resources, including 52.8% who had ‘enough with a little extra’ and 30.6% who ‘always had money left over.’ Care recipients of these care partners had a mean age of 82.0 years (SD = 8.5); 47.2% had Alzheimer’s disease, 83.3% had moderate dementia, and 77.8% lived with the care partner. Care partner AE status was not associated with study withdrawal (Fisher’s exact p = 0.852; adjusted OR = 0.86, 95% CI 0.38-1.90, p = 0.720).

**Table 1.** Baseline characteristics by care partner adverse event status.

| **Characteristic** | **No AE (N=152)** | **AE (N=36)** |
|---|---|---|
| Age (Person with Dementia, years) |  |  |
| Mean (SD) | 80.5 (8.17) | 81.9 (8.64) |
| Median [Min, Max] | 80.5 [56.0, 100] | 83.5 [61.0, 94.0] |
| Missing | 2 (1.3%) | 0 (0%) |
| Race (Person with Dementia) |  |  |
| American Indian or Alaska Native | 1 (0.7%) | 0 (0%) |
| Asian | 4 (2.6%) | 0 (0%) |
| Black | 11 (7.2%) | 4 (11.1%) |
| Caucasian | 130 (85.5%) | 32 (88.9%) |
| Native Hawaiian or Pacific Islander | 1 (0.7%) | 0 (0%) |
| Other | 4 (2.6%) | 0 (0%) |
| Missing | 1 (0.7%) | 0 (0%) |
| Hispanic ethnicity (Person with Dementia) |  |  |
| No | 141 (92.8%) | 35 (97.2%) |
| Yes | 10 (6.6%) | 1 (2.8%) |
| Missing | 1 (0.7%) | 0 (0%) |
| Gender (Person with Dementia) |  |  |
| Female | 85 (55.9%) | 21 (58.3%) |
| Male | 66 (43.4%) | 15 (41.7%) |
| Missing | 1 (0.7%) | 0 (0%) |
| Sexual orientation (Person with Dementia) |  |  |
| Bisexual | 4 (2.6%) | 0 (0%) |
| Gay | 1 (0.7%) | 1 (2.8%) |
| Heterosexual | 144 (94.7%) | 34 (94.4%) |
| Prefer not to answer/Missing | 3 (2.0%) | 0 (0%) |
| Lesbian | 0 (0%) | 1 (2.8%) |
| Education (Person with Dementia) |  |  |
| Mean (SD) | 15.4 (2.75) | 15.5 (2.82) |
| Median [Min, Max] | 16.0 [8.00, 20.0] | 16.0 [12.0, 20.0] |
| Missing | 1 (0.7%) | 0 (0%) |
| Dementia diagnosis (Person with Dementia) |  |  |
| Alzheimer's | 62 (40.8%) | 17 (47.2%) |
| FTD (PPA/Behavioral Variant) | 15 (9.9%) | 2 (5.6%) |
| Lewy Body | 10 (6.6%) | 3 (8.3%) |
| Mixed | 12 (7.9%) | 4 (11.1%) |
| Vascular | 13 (8.6%) | 2 (5.6%) |
| Other/Don't know | 40 (26.3%) | 8 (22.2%) |
| Dementia stage (Person with Dementia) |  |  |
| Moderate | 134 (88.2%) | 30 (83.3%) |
| Severe | 18 (11.8%) | 6 (16.7%) |
| **Lives with care partner** |  |  |
| Does not live with Care Partner | 41 (27.0%) | 8 (22.2%) |
| Lives with Care Partner | 110 (72.4%) | 28 (77.8%) |
| Missing | 1 (0.7%) | 0 (0%) |
| **Care partner age (years)** |  |  |
| Mean (SD) | 66.7 (12.4) | 68.7 (9.86) |
| Median [Min, Max] | 67.0 [32.0, 91.0] | 69.0 [50.0, 86.0] |
| **Care partner race** |  |  |
| Asian | 4 (2.6%) | 0 (0%) |
| Black | 11 (7.2%) | 3 (8.3%) |
| Caucasian | 128 (84.2%) | 33 (91.7%) |
| Native Hawaiian or Pacific Islander | 2 (1.3%) | 0 (0%) |
| Other | 7 (4.6%) | 0 (0%) |
| **Care partner Hispanic ethnicity** |  |  |
| No | 143 (94.1%) | 35 (97.2%) |
| Yes | 9 (5.9%) | 1 (2.8%) |
| **Rural status** |  |  |
| Rural | 21 (13.8%) | 7 (19.4%) |
| Urban | 125 (82.2%) | 29 (80.6%) |
| Missing | 6 (3.9%) | 0 (0%) |
| **Care partner gender** |  |  |
| Female | 112 (73.7%) | 26 (72.2%) |
| Male | 38 (25.0%) | 10 (27.8%) |
| Missing | 2 (1.3%) | 0 (0%) |
| **Care partner sexual orientation** |  |  |
| Bisexual | 4 (2.6%) | 1 (2.8%) |
| Gay | 1 (0.7%) | 2 (5.6%) |
| Heterosexual | 140 (92.1%) | 31 (86.1%) |
| Lesbian | 2 (1.3%) | 2 (5.6%) |
| Other | 2 (1.3%) | 0 (0%) |
| Prefer not to answer/Missing | 3 (2.0%) | 0 (0%) |
| **Care partner education** |  |  |
| Mean (SD) | 16.3 (2.20) | 16.2 (2.06) |
| Median [Min, Max] | 16.0 [12.0, 20.0] | 16.0 [12.0, 20.0] |
| **Years caregiving** |  |  |
| Mean (SD) | 4.39 (3.41) | 3.89 (3.87) |
| Median [Min, Max] | 3.25 [1.00, 23.0] | 3.00 [1.00, 21.0] |
| **Relationship to person with dementia** |  |  |
| Child | 61 (40.1%) | 14 (38.9%) |
| Other | 2 (1.3%) | 1 (2.8%) |
| Other relative | 2 (1.3%) | 0 (0%) |
| Sibling | 3 (2.0%) | 1 (2.8%) |
| Spouse/Partner | 84 (55.3%) | 20 (55.6%) |
| **Income sufficiency** |  |  |
| I always have more left over | 46 (30.3%) | 11 (30.6%) |
| I can't make ends meet | 4 (2.6%) | 0 (0%) |
| I have enough with a little extra | 73 (48.0%) | 19 (52.8%) |
| I have just enough | 29 (19.1%) | 6 (16.7%) |
| **Baseline total RMBPC reactivity** |  |  |
| Mean (SD) | 40.4 (15.2) | 42.0 (16.0) |
| Median [Min, Max] | 40.5 [0, 81.6] | 45.7 [0, 64.0] |
| Missing | 11 (7.2%) | 1 (2.8%) |
| **Baseline MM-CGI grief score** |  |  |
| Mean (SD) | 57.8 (12.0) | 57.6 (14.1) |
| Median [Min, Max] | 58.0 [18.0, 89.0] | 59.0 [29.0, 84.0] |
| Missing | 12 (7.9%) | 2 (5.6%) |
| **Baseline CES-D total score** |  |  |
| Mean (SD) | 11.9 (6.05) | 12.1 (6.67) |
| Median [Min, Max] | 11.0 [0, 27.0] | 12.0 [1.00, 28.0] |
| Missing | 12 (7.9%) | 2 (5.6%) |

Note. No statistically significant differences were observed between groups for baseline care partner demographics, caregiving characteristics, person with dementia characteristics, baseline reactivity, depressive symptoms, or grief (all p > .05).

### ADVERSE EVENTS

Forty-three care partner AEs were reported by 36 care partners during Tele-STELLA. Most care partners experienced one AE (29/36, 80.6%), while seven (19.4%) experienced two AEs. One care partner AE was classified as severe. Physical AEs accounted for 48.8% (21/43) of events, Psychological AEs for 39.5% (17/43), and Random/Other AEs for 11.6% (5/43) (Figure 1). No aggression-related AE in the available dataset documented a care partner injury or medical visit directly attributable to care-recipient aggression.

**Figure 1.**
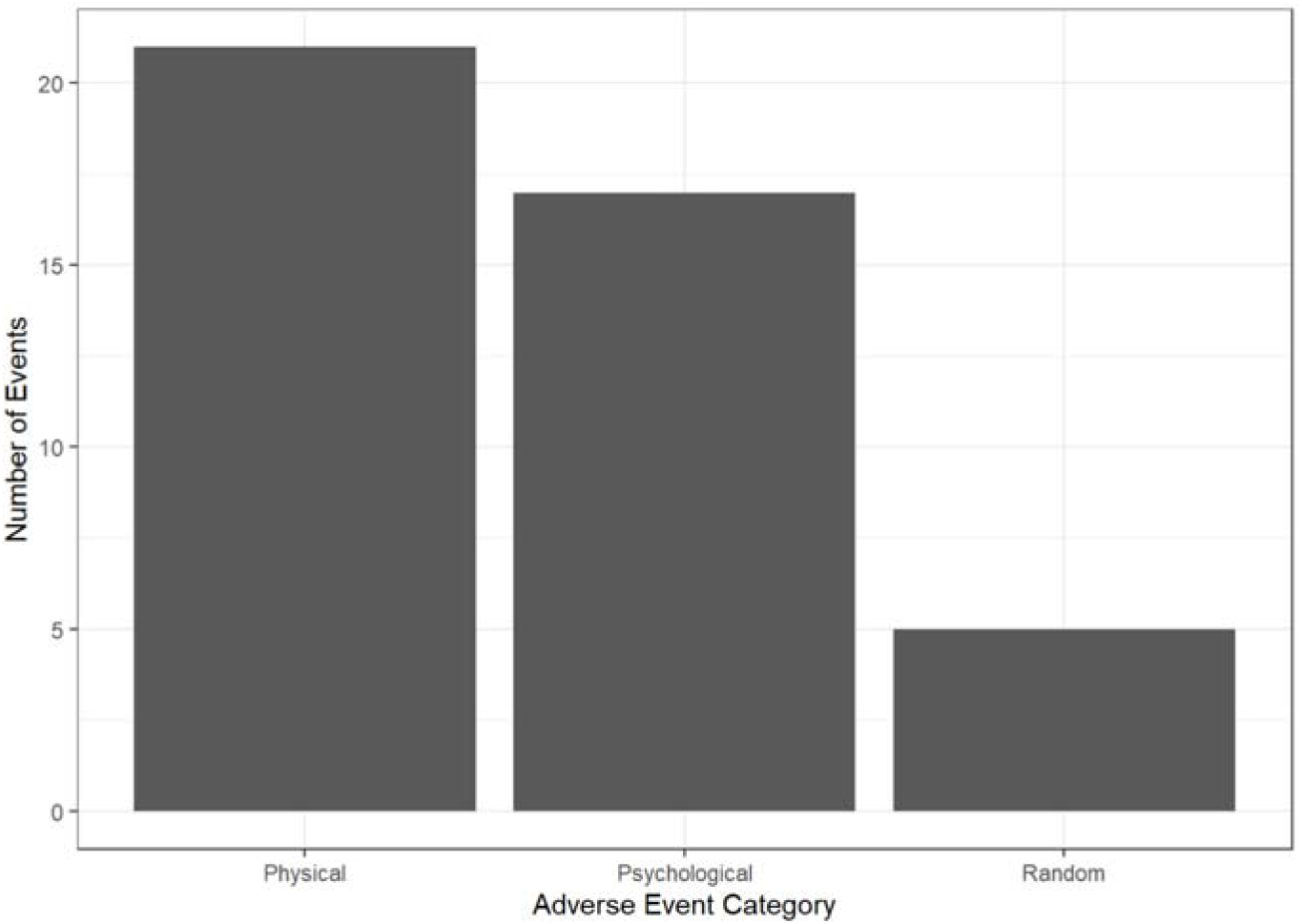
Distribution of adverse event types.

Among Physical AEs, infections (e.g., flu infections) were most common (13/21, 61.9%), followed by chronic disease exacerbations (5/21, 23.8%), pain associated with physical care (2/21, 9.5%), and falls (1/21, 4.8%). Psychological AEs were evenly divided between stress/distress associated with care partner (8/17, 47.1%) and aggression from the care recipient (8/17, 47.1%); one additional event was classified as Other Psychological (1/17, 5.9%). Random/Other events included motor vehicle accidents (2/5, 40.0%), dog aggression (2/5, 40.0%), and being struck by a vehicle (1/5, 20.0%). AE subcategories are summarized in Table 2.

**Table 2.**
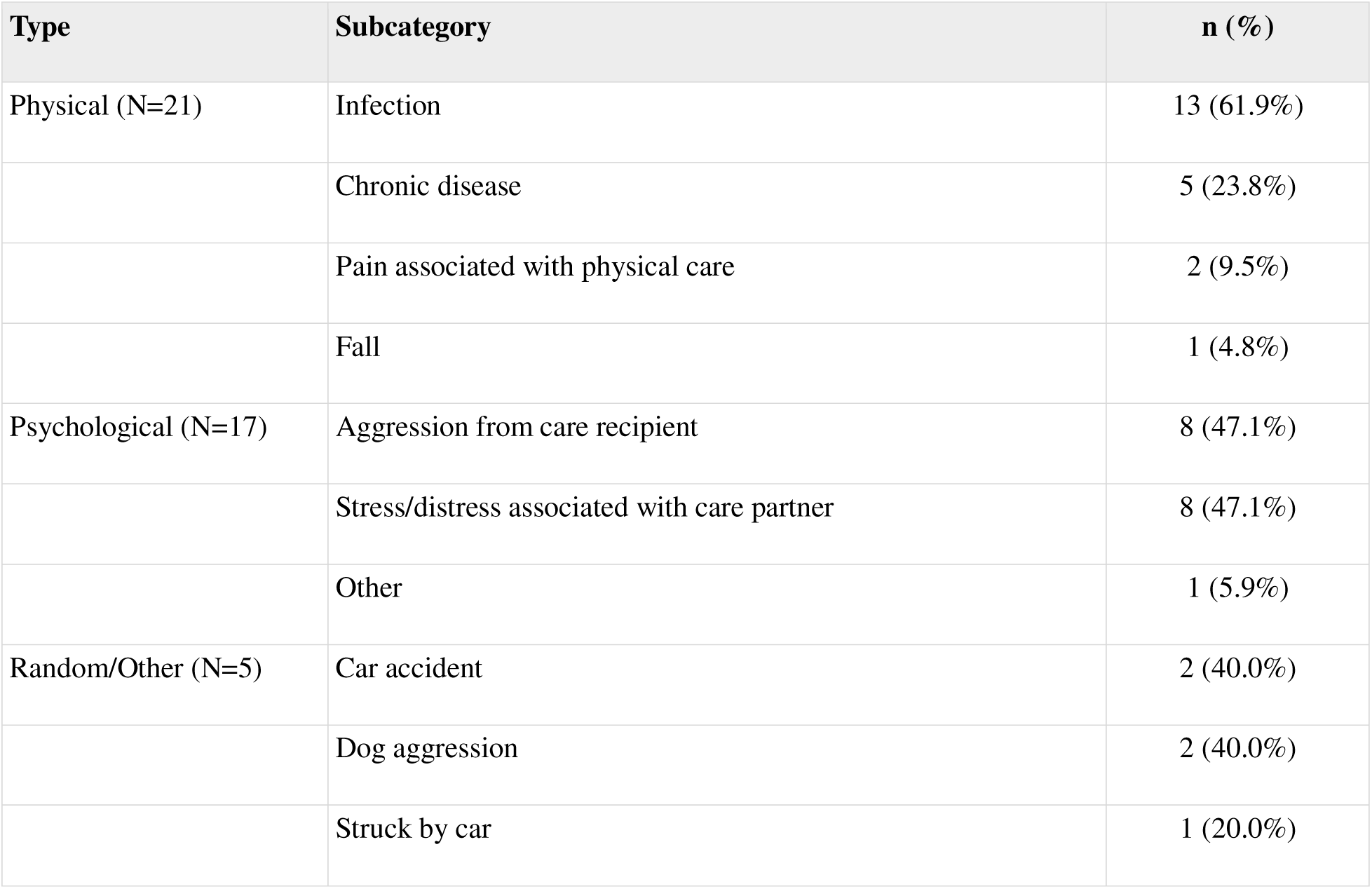
Subcategories of adverse event types.

| **Type** | **Subcategory** | **n (%)** |
|---|---|---|
| Physical (N=21) | Infection | 13 (61.9%) |
|  | Chronic disease | 5 (23.8%) |
|  | Pain associated with physical care | 2 (9.5%) |
|  | Fall | 1 (4.8%) |
| Psychological (N=17) | Aggression from care recipient | 8 (47.1%) |
|  | Stress/distress associated with care partner | 8 (47.1%) |
|  | Other | 1 (5.9%) |
| Random/Other (N=5) | Car accident | 2 (40.0%) |
|  | Dog aggression | 2 (40.0%) |
|  | Struck by car | 1 (20.0%) |

### BASELINE PSYCHOLOGICAL DISTRESS

Among care partners with AEs, mean baseline RMBPC reactivity was 37.2 (SD = 20.1), MM-CGI grief was 58.3 (SD = 14.7), and CES-D depressive symptoms were 11.7 (SD = 7.0), with substantial variability across participants (Figure 2).

**Figure 2.**
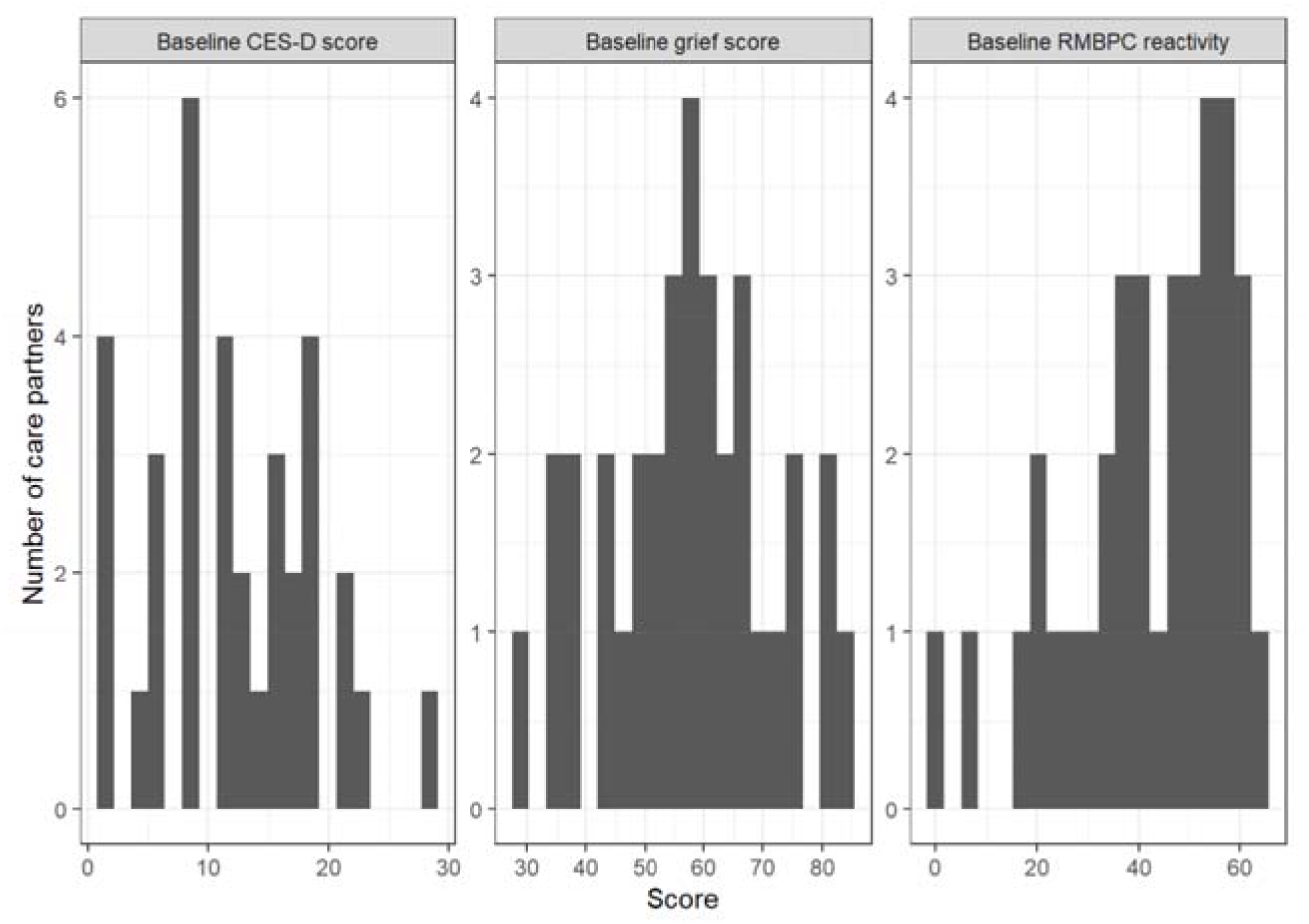
Distribution of baseline care partner burden and psychological distress measures.

### BASELINE BURDEN BY ADVERSE EVENT CATEGORY

Exploratory category-level comparisons showed more consistent differences for Psychological than for Physical AEs. Care partners who experienced a Psychological AE had higher baseline grief (MM-CGI: 65.1 vs. 53.5; mean difference = –11.58, 95% CI –20.03 to –3.14; Hedges’ g = –0.86; p = 0.009) and higher RMBPC reactivity (49.3 vs. 37.7; mean difference = –11.55, 95% CI-21.12 to –1.97; g = –0.74; p = 0.020) than care partners who did not experience a Psychological AE. Conversely, care partners who experienced a Physical AE had lower RMBPC reactivity than care partners who did not experience a Physical AE (36.8 vs. 49.0; mean difference = 12.18, 95% CI 2.45 to 21.91; g = 0.79; p = 0.016). Baseline depressive symptoms did not differ significantly by overall AE category. Category-level comparisons are presented in Table 3.

**Table 3.** Baseline psychological measures by adverse event category.

| **Outcome** | **AE category** | **Category absent, n / Mean (SD)** | **Category present, n / Mean (SD)** | **Mean difference (95% CI)** | **Hedges' g** | **p-value** |
|---|---|---|---|---|---|---|
| CES-D | Other | 31 / 12.1 (7.0) | 3 / 13.0 (1.0) | -0.94 (-3.77, 1.90) | -0.13 | 0.504 |
| CES-D | Physical | 14 / 14.2 (6.6) | 20 / 10.7 (6.5) | 3.51 (-1.15, 8.18) | 0.53 | 0.134 |
| CES-D | Psychological | 22 / 10.9 (6.2) | 12 / 14.5 (7.1) | -3.64 (-8.71, 1.44) | -0.54 | 0.151 |
| MM-CGI | Other | 31 / 58.4 (14.2) | 3 / 49.3 (13.4) | 9.05 (-20.53, 38.64) | 0.63 | 0.364 |
| MM-CGI | Physical | 14 / 62.1 (12.1) | 20 / 54.4 (14.9) | 7.74 (-1.71, 17.19) | 0.55 | 0.105 |
| MM-CGI | Psychological | 22 / 53.5 (14.8) | 12 / 65.1 (9.3) | -11.58 (-20.03, -3.14) | -0.86 | 0.009 |
| RMBPC reactivity | Other | 32 / 41.2 (16.3) | 3 / 50.3 (10.4) | -9.13 (-30.17, 11.90) | -0.56 | 0.262 |
| RMBPC reactivity | Physical | 15 / 49.0 (10.3) | 20 / 36.8 (17.7) | 12.18 (2.45, 21.91) | 0.79 | 0.016 |
| RMBPC reactivity | Psychological | 22 / 37.7 (17.3) | 13 / 49.3 (10.5) | -11.55 (-21.12, -1.97) | -0.74 | 0.020 |

\* Mean differences are calculated as category absent minus category present.

### BASELINE BURDEN BY ADVERSE EVENT SUBCATEGORY

At the subcategory level, care partners who experienced a stress/distress-related AE had substantially higher baseline depressive symptoms (18.3 vs. 10.8; mean difference = –7.51, 95% CI –14.06 to –0.96; g = –1.20; p = 0.030) and higher RMBPC reactivity (55.4 vs. 38.6; mean difference = –16.75, 95% CI –26.17 to –7.33; g = –1.11; p = 0.002) than care partners without these events. Aggression-related AEs were associated with higher grief scores (64.2 vs. 56.4; mean difference = –7.75, 95% CI –14.67 to –0.83; g = –0.54; p = 0.030), whereas infection-related AEs were associated with lower grief scores (49.8 vs. 61.8; mean difference = 11.98, 95% CI 2.25 to 21.72; g = 0.89; p = 0.018). Estimates for other subcategories should be interpreted cautiously because of small sample sizes. Subcategory-level comparisons are presented in Table 4.

**Table 4.** Baseline psychological measures by adverse event subcategory.

| **Outcome** | **Subcategory** | **Subcategory absent, n / Mean (SD)** | **Subcategory present, n / Mean (SD)** | **Mean difference (95% CI)** | **Hedges' g** | **p-value** |
|---|---|---|---|---|---|---|
| CES-D | Aggression from care recipient | 29 / 12.1 (7.0) | 5 / 12.4 (4.8) | -0.30 (-6.15, 5.56) | -0.04 | 0.909 |
| CES-D | Chronic disease | 29 / 12.3 (7.0) | 5 / 11.2 (5.0) | 1.11 (-4.96, 7.18) | 0.16 | 0.679 |
| CES-D | Infection | 22 / 13.7 (6.5) | 12 / 9.3 (6.2) | 4.35 (-0.35, 9.05) | 0.66 | 0.068 |
| CES-D | Stress/distress associated with care partner | 28 / 10.8 (6.1) | 6 / 18.3 (6.2) | -7.51 (-14.06, -0.96) | -1.20 | 0.030 |
| MM-CGI | Aggression from care recipient | 29 / 56.4 (15.0) | 5 / 64.2 (4.2) | -7.75 (-14.67, -0.83) | -0.54 | 0.030 |
| MM-CGI | Chronic disease | 29 / 57.2 (14.7) | 5 / 59.6 (11.6) | -2.36 (-16.50, 11.78) | -0.16 | 0.701 |
| MM-CGI | Infection | 22 / 61.8 (13.1) | 12 / 49.8 (13.1) | 11.98 (2.25, 21.72) | 0.89 | 0.018 |
| MM-CGI | Stress/distress associated with care partner | 28 / 55.9 (14.0) | 6 / 65.3 (13.2) | -9.40 (-23.33, 4.52) | -0.66 | 0.157 |
| RMBPC reactivity | Aggression from care recipient | 30 / 41.6 (17.2) | 5 / 44.3 (5.8) | -2.73 (-11.29, 5.83) | -0.16 | 0.512 |
| RMBPC reactivity | Chronic disease | 30 / 42.3 (16.2) | 5 / 40.3 (16.6) | 1.99 (-18.15, 22.12) | 0.12 | 0.813 |
| RMBPC reactivity | Infection | 23 / 44.7 (15.0) | 12 / 36.8 (17.3) | 7.89 (-4.39, 20.17) | 0.49 | 0.195 |
| RMBPC reactivity | Stress/distress associated with care partner | 28 / 38.6 (15.7) | 7 / 55.4 (8.8) | -16.75 (-26.17, -7.33) | -1.11 | 0.002 |

\* Subcategories with fewer than three care partners experiencing the event were omitted because estimates were unstable. Mean differences are calculated as subcategory absent minus subcategory present.

## DISCUSSION & IMPLICATIONS

This analysis provides an initial characterization of care partner AEs reported during the Tele-STELLA study and demonstrates that AE narratives can capture more than conventional medical occurrences. Among 43 care partner AEs reported by 36 care partners during Tele-STELLA, approximately half were Physical, nearly 40% were Psychological, and the remainder were Random/Other events. Physical events were dominated by infections and chronic disease exacerbations, whereas Psychological events were almost evenly divided between care partner stress/distress and aggression from the care recipient. Exploratory analyses further suggested that Psychological AEs occurred within a different baseline care partner context than Physical AEs: care partners who experienced Psychological AEs had greater grief and reactivity to dementia-related behavioral symptoms, stress/distress-related AEs were characterized by higher depressive symptoms and behavioral reactivity, and aggression-related AEs were characterized by greater grief. Together, these findings suggest that classifying care partner AEs according to their clinical and care partner context may reveal information that is obscured when all AEs are treated as a single outcome.

The high proportion of Physical AEs is consistent with a broader literature demonstrating that caring for a person with dementia can affect care partners’ own health. Family care partners may prioritize the needs of the person living with dementia while delaying or reducing attention to their own health, and being a care partner has been associated with poorer physical health and reduced engagement in health-promoting behaviors.^1,2,7,24^ Infections and chronic disease exacerbations accounted for most Physical AEs in the present analysis, reinforcing the importance of viewing the care partner as a member of the clinical dyad rather than solely as a source of support. At the same time, care partners with Physical AEs had lower baseline RMBPC reactivity, and those with infection-related AEs had lower grief scores. These findings should not be interpreted as evidence that lower psychological burden increases vulnerability to physical illness. Rather, the contrasting baseline profiles suggest that Physical and Psychological AEs may reflect different aspects of care partner vulnerability and warrant separate examination in future research.

Psychological AEs may be particularly informative for understanding heterogeneity in care partners’ adverse experiences. Dementia care partnering is characterized by substantial emotional demands, including depression, anxiety, grief, isolation, and chronic stress, and behavioral and psychological symptoms of dementia are repeatedly identified as major contributors to care partner burden.^1,2,6,14^ In the present analysis, care partners who experienced Psychological AEs had greater baseline grief and RMBPC reactivity. Depressive symptoms did not differ significantly by the broader Psychological AE category, but a more differentiated pattern emerged when events were separated into subcategories: stress/distress-related AEs were associated with substantially higher depressive symptoms and behavioral reactivity, whereas aggression-related AEs were associated with greater grief. This pattern suggests that different dimensions of care partner psychological burden may be relevant to different forms of adverse experience rather than representing a single generalized construct.

The relationship with RMBPC reactivity may be particularly important. Care partners who experienced Psychological AEs reported greater baseline reactivity to dementia-related behavioral symptoms than those who did not experience a Psychological AE. This finding highlights care partner appraisal and response to behavioral symptoms as a potentially important dimension of psychological vulnerability.

Prior work similarly emphasizes the importance of care partner confidence, appraisal, and perceived ability to manage challenging behaviors.^14,15^ Intervention studies provide convergent support: TeleFAMILIES improved care partner RMBPC reaction scores,^25^ improvements in care partner preparedness during ICECaP were associated with reductions in negative reactions to behavioral symptoms,^26^ and virtual care partner education increased confidence, self-efficacy, and perceived ability to manage dementia-related behaviors.^27^ These findings support moving beyond inventories of which behaviors are present toward greater attention to how care partners experience, interpret, and manage them.

The association between aggression-related AEs and greater care partner grief provides an additional window into the relational context of care partner. Aggression is among the most difficult dementia-related behaviors for family care partners to manage and may include verbal threats, physical aggression, property destruction, and other behaviors that place care partners at physical or emotional risk.^11,14,15,17,18^ Previous studies describe depression, anxiety, anger, isolation, helplessness, and substantial emotional distress among family care partners exposed to violent or harmful behavior from care recipients.^6,14,15,17^ In the present dataset, no aggression-related AE narrative documented a care partner injury or medical visit directly attributable to the aggressive event, although the available narratives varied from threatened physical harm to nonspecific reports of abuse. The clinical significance of aggression therefore should not be limited to episodes that culminate in documented physical injury. Dementia care partnering is also accompanied by ongoing losses in relationship, role, communication, and the identity of the person living with dementia, making pre-death grief an important component of the care partner experience.^2,9,19,28^ Greater grief among care partners experiencing aggression-related AEs may reflect a particularly difficult intersection between relational loss and behavioral change. An aggressive episode occurs within a relationship that often predates dementia by decades and may be experienced against a backdrop of profound change in that relationship. These findings may also have clinical relevance for the broader care partner dyad. Tele-STELLA is informed by Kales et al.’s framework, which emphasizes the multidirectional relationship between care partner and person-with-dementia well-being.^29^

Care partner stress and behavioral and psychological symptoms of dementia have been linked with healthcare utilization, hospitalization, institutionalization, and poorer quality of life for both members of the dyad.^6,14,15,28^ Emerging telehealth research further suggests that care partner– and dyad-focused interventions can affect both care partner responses and downstream healthcare outcomes. In a virtual iSupport trial, care partners demonstrated improved self-efficacy, social support, and reactions to behavior, while care recipients had a lower hospital admission rate.^30^ A telehealth-facilitated palliative care intervention similarly reported fewer emergency department visits and hospitalizations among people living with dementia.^31^ These intervention findings do not establish that care partner AEs or psychological burden cause healthcare utilization. They do, however, support studying care partner experiences, care-recipient outcomes, and healthcare events together when evaluating periods of dyadic vulnerability.

A natural next step is to examine how care partner strain and AEs change over time and whether changes in one precede changes in the other. Periods of increased emotional or physical strain may precede an AE, while experiencing an AE may itself increase subsequent strain. Repeated longitudinal assessments could distinguish relatively stable differences between care partners from shorter-term changes within an individual and clarify whether the relationship between strain and AEs is potentially bidirectional. Technology-enabled care partner studies demonstrate the feasibility of collecting repeated data remotely, including daily ecological momentary assessments of care partnerwell-being,^23,32,30^ and longer-term remote monitoring within the care partner environment.^33^ Aligning repeated strain measures with the timing of care partner and care-recipient AEs could therefore provide a more dynamic picture of vulnerability than baseline measures alone. Future work should also examine whether care partner and care-recipient AEs, alone or in combination with changes in care partner strain, are associated with hospitalization, emergency healthcare use, transitions in living situation, or institutionalization.

This line of research may ultimately identify modifiable targets for care partner interventions. The present findings do not support using grief, depressive symptoms, or behavioral reactivity as clinical predictors of AEs; rather, they identify constructs that warrant prospective study. Importantly, several of these domains appear responsive to intervention. Tele-Savvy produced improvements in depression, perceived stress, reactions to care-recipient behaviors, and care partner mastery.^20^ TeleFAMILIES was associated with reductions in care partner burden and depressive symptoms as well as improved RMBPC reaction scores.^25^ ICECaP improved care partner preparedness, and greater improvement in preparedness was associated with reductions in burden and negative reactions to behavioral symptoms of dementia.^26^ Virtual care partner education has also improved care partner confidence and self-efficacy,^27^ while a videoconference-delivered acceptance and commitment therapy program showed improvements in stress, psychological quality of life, and several within-group psychological outcomes, including pre-death grief.^34^ Taken together, these findings reinforce the possibility that changing the caregiver’s appraisal, preparedness, and response to difficult circumstances may be meaningful even when the underlying dementia or behavioral symptom cannot be eliminated. This analysis also extends our group’s prior work using telehealth to understand and address care partner responses to dementia-related behavioral symptoms.^28,35^ Whereas earlier studies focused primarily on care partner burden, behavioral reactivity, and intervention outcomes, the present analysis examines care partner AEs as an additional source of information about periods of potential vulnerability within the care partner dyad.^19,28,35^

More broadly, the present findings support conceptualizing AEs as part of the lived trajectory of dementia care partner rather than solely as regulatory occurrences recorded during research participation. Care partner AEs have received relatively little attention as analytic outcomes in care partner and dyadic studies. In at least some recent care partner intervention reports, safety information is summarized at the level of intervention-related harm; for example, one mHealth care partner trial reported that no intervention-related AEs occurred,^36^ rather than characterizing the broader events experienced by care partners during participation. Tele-STELLA’s systematic collection of care partner AEs created an opportunity to examine these reports as a source of clinically meaningful information. Narrative AE reports may capture periods in which the care partner system is under unusual physical or psychological strain and systematically classifying these events may offer an underused approach to understanding care partner health, behavioral challenges, and points of vulnerability over time. Combining AE surveillance with repeated measures of emotional strain, physical strain, grief, behavioral reactivity, and clinically meaningful dyadic outcomes may reveal patterns that are not apparent from conventional cross-sectional assessments.

This analysis has several strengths. AEs were collected during care partners’ participation in a longitudinal telehealth intervention and were available as free-text narratives, allowing events to be examined beyond a simple count of safety occurrences. The sample included care partners ranging from 50 to 86 years of age, both urban and rural participants, and care recipients with several dementia diagnoses, including Alzheimer’s disease, Lewy body dementia, mixed and vascular dementia, and frontotemporal dementia/primary progressive aphasia. Linking AE narratives with baseline measures of grief, depressive symptoms, and behavioral reactivity also allowed the descriptive classification to be considered within a broader care partner context.

Several limitations should be considered. Although the overall Tele-STELLA cohort was larger, the AE-specific analyses were based on 36 care partners who experienced at least one reported AE, limiting precision for category– and subcategory-level comparisons. The cohort was also demographically homogeneous, with most care partners identifying as White and female, which limits generalizability. Category– and subcategory-level comparisons were exploratory, involved small groups and missing data on some measures, and included numerous statistical comparisons. The observed effect sizes and p-values should therefore be interpreted as hypothesis-generating rather than confirmatory.

The AE narratives themselves also impose important limitations. Reporting depended on care partners and study staff recognizing and documenting an event, and some events may have gone unreported because they were perceived as minor, were forgotten, or involved information a participant was reluctant to disclose. Narrative detail varied considerably, requiring investigator judgment during classification and limiting the ability to distinguish features such as verbal versus physical aggression or the presence of undocumented minor injury. Although no aggression-related AE documented a care partner injury or medical visit directly attributable to aggression, absence of documentation should not be interpreted as evidence that physical harm never occurred. Finally, baseline psychological characteristics were compared with whether care partners experienced each AE type at any point during study participation; these associations do not establish causal mechanisms or demonstrate that baseline grief, depression, or reactivity predicts future AEs.

In conclusion, care partner AEs reported during the Tele-STELLA study comprised a heterogeneous mixture of physical illness, care partner stress and distress, aggression from the person living with dementia, and unrelated external events. Psychological AEs were characterized by greater baseline grief and reactivity to dementia-related behaviors; within this category, stress/distress-related AEs were associated with depressive symptoms and behavioral reactivity, whereas aggression-related AEs were associated with greater grief. These findings suggest that understanding how care partners experience and respond to the psychological and behavioral demands of dementia may be important for interpreting AEs within care partner dyads. Prospective studies that align repeated measures of care partner strain with care partner and care-recipient AEs and downstream healthcare or residential outcomes are an important next step toward identifying periods of vulnerability and developing timely interventions that support both members of the dyad.

## Acknowledgments

We thank the care partners and their family members who participated in Tele-STELLA and the study staff who supported participant follow-up and adverse event documentation.

## Author contributions

Conception or design of the work: C. McClure, A. Lindauer. Acquisition of data: H. Chatterton. Analysis of data: H. Franklin. Interpretation of data: C. McClure, H. Franklin, A. Lindauer. Drafting of the manuscript: C. McClure, H. Chatterton, H. Franklin, A. Lindauer. Revision of the manuscript: C. McClure, H. Chatterton, H. Franklin, A. Mooney, A. Lindauer.

## STATEMENTS AND DECLARATIONS

### Ethical Considerations

The Tele-STELLA study was approved by the Oregon Health & Science University Institutional Review Board (IRB #: 22288).

### Consent to Participate

All participants provide written informed consent prior to participating in this study.

### Consent for Publication

Not applicable.

### Data Availability

The data supporting the findings of this study are available from the Oregon Life Laboratory within the Digital Technology Core (DTC) of the Oregon Alzheimer’s Disease Research Center (OADRC) and may be requested at: https://octri.ohsu.edu/redcap/surveys/?s=EW4HKW47XH&_ga=2.9914869.1166959164.1653610093-1864845509.1632940067

